# Mandatory or Voluntary COVID-19 Vaccination: Insights into the Knowledge, Attitude and Perception among Healthcare Workers at a Nursing College in South Africa

**DOI:** 10.1101/2024.08.20.24312309

**Authors:** Lindokuhle Mokoena, Tanusha Singh

## Abstract

Vaccine hesitancy has emerged as a significant global challenge impacting healthcare institutions, workplaces and governments alike. Despite concerted efforts by the government and numerous other institutions in South Africa, low vaccination rates persist (33% as of November 13, 2023), reflecting the persistence of this global challenge. This challenge is particularly pronounced in educational institutions such as institutions of higher learning in South Africa, where many people converge, increasing the risk of viral exposure. This study aims to assess the knowledge, attitudes, and perceptions of healthcare workers (HCWs) at a Nursing College regarding voluntary and mandatory COVID-19 vaccination. Employing a quantitative approach, a survey with closed-ended Likert-type questions was administered to 504 individuals at the College. The sample size of 218 respondents was calculated based on a margin of error of 5%, a confidence level of 95%, and an additional 25% contingency for potential incomplete data, resulting in a final representative sample of 270 respondents. The Statistical Package for Social Sciences (SPSS) was used for the analysis. Findings indicate a high uptake of voluntary COVID-19 vaccinations among HCWs, with some being mandated. Most HCWs demonstrated a strong commitment to safeguarding themselves and others. Despite concerns, HCWs thought the COVID-19 vaccines were effective, and their views were supported by a low level of infection among the participants, underscoring its efficacy in preventing transmission. Effective communication emerged as a critical factor in addressing post-vaccination behaviours and enhancing vaccine acceptance. However, the findings also highlighted the need for tailored outreach strategies to specific audiences, such as pregnant women, and the importance of addressing concerns about adverse effects through clear and open communication. Several factors influencing the choice between mandatory and voluntary vaccination were identified, including eligibility concerns, religious convictions, and financial considerations. Notably, concerns about safety and knowledge gaps outweighed these factors, suggesting the need for targeted educational initiatives to bolster vaccine acceptance. In conclusion, this study provides valuable insights into the dynamics of vaccination acceptance among an influential occupational group, with implications for the acceptance of other vaccines. Vaccination efforts can be strengthened by addressing concerns, enhancing communication strategies, and tailoring outreach efforts to promote public health in light of future outbreaks.

**Author summary:** Conception and design of the study: LM, TS

Data acquisition: LM

Data analysis: LM

Interpretation of the data: LM, TS

Drafting of the paper: LM

Critical revision of the paper: TS

Both authors approved the final manuscript

## Introduction

The evaluation of healthcare workers’ (HCWs) views on voluntary and mandatory COVID-19 vaccination exposes various factors that impact their decision to get vaccinated. Research suggests that a notable portion of HCWs have an understanding of COVID-19 vaccines, which is a key determinant in their willingness to be vaccinated. For instance, a study conducted at a hospital in southern Nigeria showed that 76.6% of HCWs had good knowledge about COVID-19 vaccines, and 63% expressed readiness to receive the vaccine. Factors like age, sex, duration of employment and knowledge about the vaccines were found to influence their willingness.^1^ Similarly, in Ghana, over half of HCWs were knowledgeable about COVID-19 vaccination. The majority held positive views on its efficacy, as evidenced by 85.9% having received at least one vaccine dose.^2^ While HCWs generally have a positive attitude towards vaccination, initial hesitancy among some is not uncommon. The latter is corroborated by the findings of another study where 93.3% of HCWs got vaccinated, with 24.2% initially hesitant but later swayed by reliable information.^3^ In Morocco, 93.3% of healthcare professionals were inoculated, with 53.1% opting for vaccination voluntarily, highlighting the significance of acceptance.^4^

In Indonesia, the perceptions of the COVID-19 vaccine were good however, their belief in its effectiveness was contrary. This highlights the importance of implementing strategies to reduce hesitancy and build trust.^5^ The views on vaccination vary among HCWs, with some expressing worries about long-term effects and profit-driven motives. Such concerns were revealed in a Malaysian study despite a low vaccine hesitancy.^6^ Other issues flagged among the Indian population were the lack of confidence in the vaccination and fear of side effects which influenced the uptake of booster doses. Furthermore, demographic factors like age, sex and residence are significant predictors.^7^ The influence of HCWs on public attitudes towards vaccination cannot be underestimated; their acceptance and endorsement of vaccines are crucial for achieving herd immunity and ensuring healthcare services.^8^ Research from Greece indicates that training HCWs and health science students on evidence-based practices is vital for improving vaccine uptake.^9^ While the level of knowledge and positive attitudes towards COVID-19 vaccination in report studies is promising, understanding the knowledge, attitudes and perceptions in a multiracial country like South Africa is important so that interventions are targeted among HCWs to address the specific issues pertaining to vaccine hesitancy. By taking this approach, healthcare workers will be able to make an impact on public health initiatives aimed at managing other emerging vaccine-related diseases. Therefore, this study aimed to assess the knowledge, attitude, and perceptions of HCWs at a Nursing College in South Africa regarding voluntary and mandatory COVID-19 vaccination. It also aimed to assess vaccination uptake, understand demographic influences, explore attitudes, and identify factors influencing the choice between mandatory and voluntary vaccination.

## Materials and methods

### Study design

This study used a quantitative cross-sectional research approach.

### Study setting

The research site is a publicly funded Nursing College situated in the Mpumalanga Province, South Africa that provides training for nurses. The respondents were employed at the research location and had convenient access to vaccination facilities. They gained valuable knowledge about the factors that may impact individuals’ decision to be vaccinated. The college’s suggestion for compulsory vaccination brought extra importance, adding to the current discussion on the choice between voluntary and mandatory immunisation.

### Study population

The study population included HCWs and nursing students enrolled at the college. The categories included the Diploma in General Nursing (R171) students, Degree in General Nursing (R425), totalling 504 individuals. Since nursing students are in the process of becoming fully qualified nurses, their involvement in patient care and the healthcare environment means they are recognised as part of the broader healthcare workforce in South Africa and thus is covered in the title by Healthcare workers. Individuals aged 18 years and above of both sexes and those who were eligible for the COVID-19 vaccination according to the National Department of Health Guidelines (SA Department of Health, 2023) met the inclusion criteria.^10^ The study adopted a stratified random sampling, ensuring that each group was represented. The sample size of 218 respondents was calculated based on a margin of error of 5% and a confidence level of 95%. Adding an additional 25% contingency for incomplete data brought the total sample size to 270 respondents, thus forming a representative sample. This sample size was allocated to each stratum based on proportional representation from the university population.

### Data collection and analysis

A closed-ended Likert-type questionnaire was utilised (S1 file). The questionnaire covered demographics, vaccination prevalence and statements related to HCWs’ knowledge, perceptions and attitudes toward voluntary and mandatory COVID-19 vaccination. The questionnaire was designed based on a comprehensive literature review and theoretical framework that identified key factors influencing COVID-19 vaccine hesitancy and acceptance. The questionnaire was piloted on ten people who were excluded from the main study. The pilot research results indicated a Cronbach’s alpha value over 0.7, confirming the consistency and reliability of the findings. Google Forms was used during the anonymised survey to collect data. A link was distributed to all students and staff randomly selected through the College email system. Follow-ups were done through email reminders three days after the questionnaire had been sent. Each respondent was expected to take 15 minutes to complete the survey. The questionnaire was administered to respondents over a period of one month, from 1 October 2023 to 31 October 2023.

Data underwent a cleaning process where incomplete surveys were eliminated and coded before being exported to the Statistical Package for Social Sciences (SPSS) (IBM, United States) for analysis. The uptake of COVID-19 vaccines among HCWs was described using measures of central tendency and measures of dispersion as of November 2023. It was also used to assess the respondents’ level of knowledge, attitude and views on voluntary and mandated COVID-19 vaccines. Inferential analyses were conducted to ascertain if there were variations in the knowledge, attitude and views of HCWs about voluntary and required COVID-19 vaccines based on socio-demographic variables. Factor analysis was used to discover the characteristics or attitudes that predict volunteer immunisations.

### Ethical statement

Ethical clearance was obtained from the University of Johannesburg’s Ethics Review Committee [ethical clearance number REC 2298-2023]. Ethical principles, including prevention of harm, confidentiality, the right to withdraw and permission-seeking, were strictly adhered to. In addition, written informed consent was obtained from participants.

## Results

### Demographic profile of respondents

The demographic profile of the 372 study respondents shows a predominance of young, single female students (table 1). The largest age group is 26-40 years, and most respondents are students. There is a significant gender imbalance, with a much higher proportion of female respondents. Marital status is mostly single, and other occupations besides students are minimally represented. This demographic profile suggests the study may be conducted in an educational setting focusing on young adult female students.

**Table 1.** Demographic characteristics of the study respondents (N = 372).

| Characteristics | Number | Percent (%) |
| --- | --- | --- |
| **Participant's Age** |  |  |
| 18 - 25 years | 75 | 20.2 |
| 26 - 40 years | 144 | 38.8 |
| 41 - 55 years | 136 | 36.7 |
| Above 55 years | 16 | 4.3 |
| **Gender of Health Workers** |  |  |
| Male | 70 | 18.8 |
| Female | 302 | 81.2 |
| **Marital status** |  |  |
| Single | 252 | 67.9 |
| Married | 103 | 27.8 |
| Divorced | 6 | 1.6 |
| Widowed | 10 | 2.7 |
| **Occupation** |  |  |
| Student | 344 | 92.5 |
| Academic staff | 21 | 5.6 |
| Admin and support staff | 7 | 1.9 |

### COVID-19 vaccination and infection status

Table 2 shows that most of the respondents surveyed were vaccinated for COVID-19, with 28.1% getting booster shots while 42.4% were vaccinated without receiving their boosters. A smaller group were awaiting their second vaccination dose. Slightly less than a quarter (23.2) had not been vaccinated. The decision to get vaccinated was mainly left to the individuals (72.0%). There were instances of infection both before and after vaccination, highlighting the risks and the importance of booster shots for added protection.

**Table 2.** COVID-19 vaccination and infection status among respondents.

| Characteristics | Number | Percent (%) |
| --- | --- | --- |
| **Vaccination status (N = 370)** |  |  |
| Fully vaccinated (including boosters) | 104 | 28.1 |
| Fully vaccinated (no boosters) | 157 | 42.4 |
| Vaccinated but awaiting 2<sup>nd</sup> dose | 23 | 6.2 |
| Not vaccinated | 86 | 23.2 |
| **Mandatory vs voluntary vaccination (N = 275)** |  |  |
| Mandatory | 77 | 28.0 |
| Voluntary | 198 | 72.0 |
| **COVID-19 infections (N = 371)** |  |  |
| Yes (before vaccination/not vaccinated) | 93 | 25.1 |
| Yes (after vaccination) | 52 | 14.0 |
| No | 226 | 60.9 |

### Respondents’ knowledge of COVID-19 vaccinations

The Likert Scale responses in Table 3 highlight a range of knowledge levels among participants regarding COVID-19 vaccinations. While 43.2% of those surveyed acknowledged the effectiveness of the vaccines, there is still scepticism, with 27.1% expressing disagreement. The majority (88.6%) recognised the importance of continuing measures post-vaccination. Opinions on the vaccine’s ability to prevent illness were split, with 57.5% in agreement and 24.8% in disagreement. The topic of implementing restrictions based on vaccination status varied, with 50.3% against and 35.1% in favour. Awareness of a vaccine information website in South Africa was high, with 80.1% agreeing with its existence. Concerning side effects, 87.6% believed there could be effects, while only 6.9% disagreed with this notion. Responses regarding vaccinating individuals with existing conditions showed mixed views, with 61.9% being supportive while 21.5% were against it. Views on vaccinating women garnered feedback, with 42.2% in favour and 39.4% opposing it. The majority (66.0%) believed that side effects typically diminish within days after vaccination; however, concerns about long-term health issues had perspectives 41.7% agreed and 30.1% disagreed.

**Table 3.**
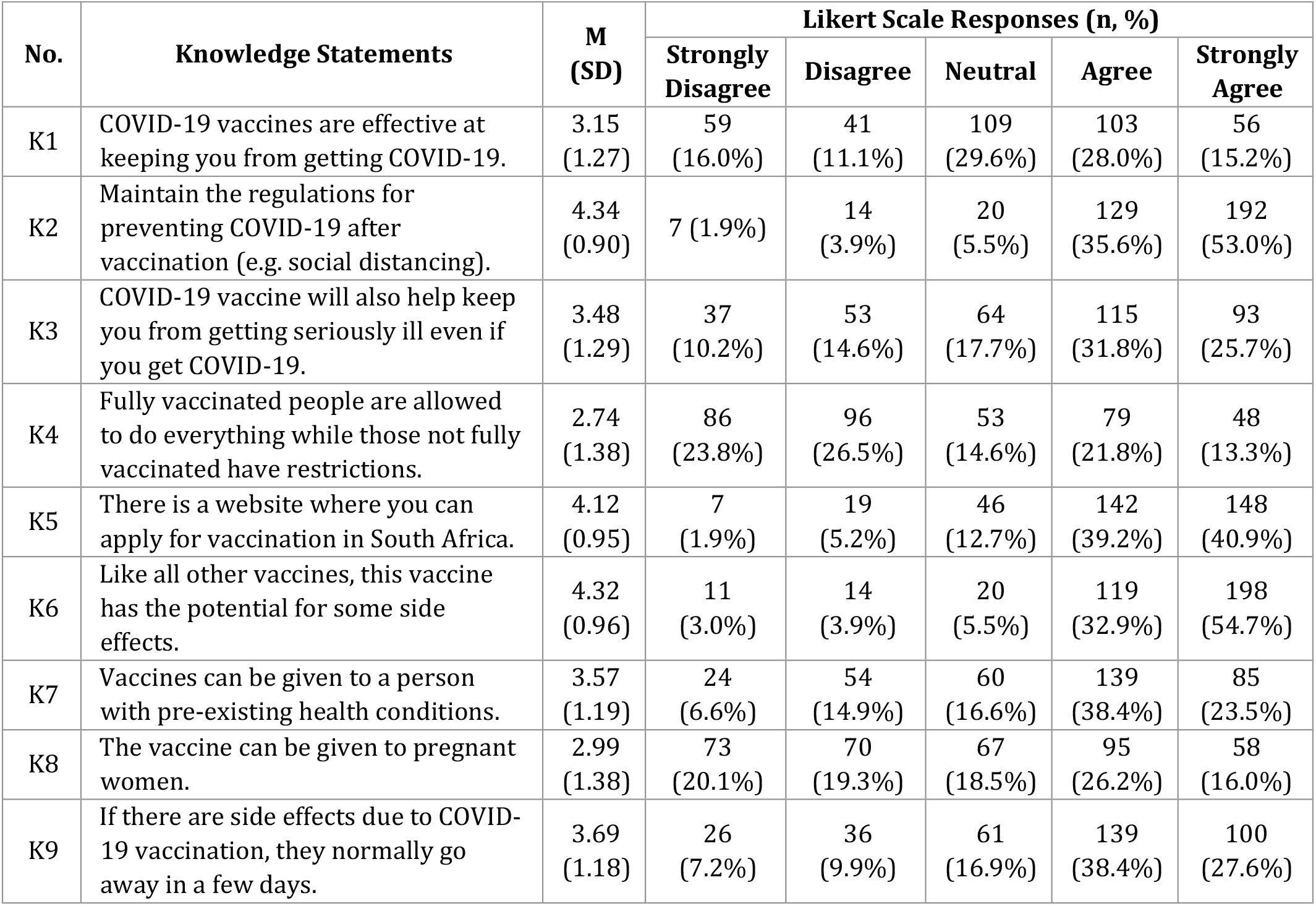
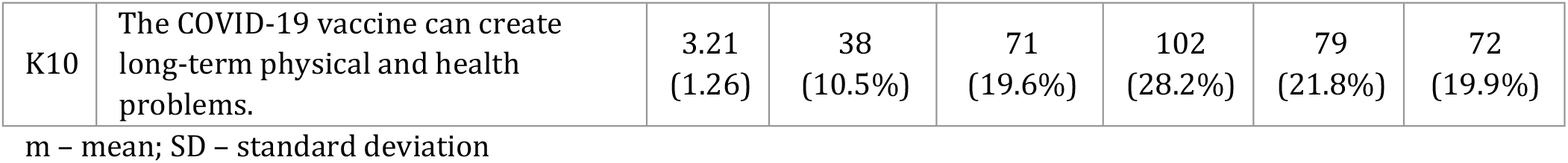
Likert Scale participant responses to knowledge on COVID-19 vaccinations (N = 372).

| No. | Knowledge Statements | M<br>(SD) | Likert Scale Responses (n, %) |  |  |  |  |
| --- | --- | --- | --- | --- | --- | --- | --- |
|  |  |  | Strongly<br>Disagree | Disagree | Neutral | Agree | Strongly<br>Agree |
| K1 | COVID-19 vaccines are effective at keeping you from getting COVID-19. | 3.15<br>(1.27) | 59<br>(16.0%) | 41<br>(11.1%) | 109<br>(29.6%) | 103<br>(28.0%) | 56<br>(15.2%) |
| K2 | Maintain the regulations for preventing COVID-19 after vaccination (e.g. social distancing). | 4.34<br>(0.90) | 7 (1.9%) | 14<br>(3.9%) | 20<br>(5.5%) | 129<br>(35.6%) | 192<br>(53.0%) |
| K3 | COVID-19 vaccine will also help keep you from getting seriously ill even if you get COVID-19. | 3.48<br>(1.29) | 37<br>(10.2%) | 53<br>(14.6%) | 64<br>(17.7%) | 115<br>(31.8%) | 93<br>(25.7%) |
| K4 | Fully vaccinated people are allowed to do everything while those not fully vaccinated have restrictions. | 2.74<br>(1.38) | 86<br>(23.8%) | 96<br>(26.5%) | 53<br>(14.6%) | 79<br>(21.8%) | 48<br>(13.3%) |
| K5 | There is a website where you can apply for vaccination in South Africa. | 4.12<br>(0.95) | 7<br>(1.9%) | 19<br>(5.2%) | 46<br>(12.7%) | 142<br>(39.2%) | 148<br>(40.9%) |
| K6 | Like all other vaccines, this vaccine has the potential for some side effects. | 4.32<br>(0.96) | 11<br>(3.0%) | 14<br>(3.9%) | 20<br>(5.5%) | 119<br>(32.9%) | 198<br>(54.7%) |
| K7 | Vaccines can be given to a person with pre-existing health conditions. | 3.57<br>(1.19) | 24<br>(6.6%) | 54<br>(14.9%) | 60<br>(16.6%) | 139<br>(38.4%) | 85<br>(23.5%) |
| K8 | The vaccine can be given to pregnant women. | 2.99<br>(1.38) | 73<br>(20.1%) | 70<br>(19.3%) | 67<br>(18.5%) | 95<br>(26.2%) | 58<br>(16.0%) |
| K9 | If there are side effects due to COVID-19 vaccination, they normally go away in a few days. | 3.69<br>(1.18) | 26<br>(7.2%) | 36<br>(9.9%) | 61<br>(16.9%) | 139<br>(38.4%) | 100<br>(27.6%) |
| K10 | The COVID-19 vaccine can create long-term physical and health problems. | 3.21<br>(1.26) | 38<br>(10.5%) | 71<br>(19.6%) | 102<br>(28.2%) | 79<br>(21.8%) | 72<br>(19.9%) |

m – mean; SD – standard deviation

### Respondents’ attitudes and perceptions towards mandatory and voluntary vaccination

The responses on the Likert Scale in Table 4 show the attitudes and perceptions regarding COVID-19 vaccinations among the participants. A significant number (86.6%) showed concern about the pandemic. In contrast, only a small percentage, 1.7%, strongly disagreed. When it comes to the safety and effectiveness of the COVID-19 vaccine, 54.2% agreed, while 14.2% disagreed. The need to get vaccinated was acknowledged by 59.2% who agreed, although some (17.0%) disagreed. There was a level of awareness that family and neighbours should receive the vaccine, with 81.2% agreeing on this point. Many also felt motivated to encourage others to get vaccinated, with 72.6% agreeing. A significant portion (68.9%) believed that vaccination could help curb the spread of COVID-19, with a minority (10.2%) disagreeing. Nearly half (49.0%) of the respondents felt uninformed about vaccines. Some hesitancy towards vaccination was observed among respondents, with one-third agreeing. Regarding opinions on mandatory vaccination for HCWs, 46.3% agreed however, slightly fewer respondents disagreed (40.3%). On the contrary, opinions on the necessity of mandatory vaccination for all South Africans varied, with fewer agreeing (36.6%) than those disagreeing (44.3%).

**Table 4.** Likert Scale participant responses regarding attitudes and perceptions towards COVID-19 vaccinations (N = 372).

| No. | Attitude and Perception Statements | M<br>(SD) | Likert Scale Responses (n, %) |  |  |  |  |
| --- | --- | --- | --- | --- | --- | --- | --- |
|  |  |  | Strongly Disagree | Disagree | Neutral | Agree | Strongly Agree |
| 1 | I am concerned about the COVID-19 pandemic. | 4.26<br>(0.85) | 6 (1.7%) | 8 (2.2%) | 34<br>(9.5%) | 149<br>(41.6%) | 161<br>(45.0%) |
| 2 | I think that the COVID-19 vaccine is safe and effective. | 3.54<br>(1.06) | 18<br>(5.0%) | 33<br>(9.2%) | 113<br>(31.6%) | 124<br>(34.6%) | 70<br>(19.6%) |
| 3 | If I am eligible for the vaccine, I need to take it as soon as possible. | 3.59<br>(1.14) | 23<br>(6.4%) | 38<br>(10.6%) | 85<br>(23.7%) | 129<br>(36.0%) | 83<br>(23.2%) |
| 4 | I am aware that my family and neighbours should take the vaccine. | 4.11<br>(0.95) | 12<br>(3.2%) | 10<br>(2.7%) | 48<br>(12.9%) | 158<br>(42.5%) | 144<br>(38.7%) |
| 5 | I should motivate my family and neighbours to take the vaccine. | 3.92<br>(1.09) | 19<br>(5.1%) | 19<br>(5.1%) | 64<br>(17.2%) | 142<br>(38.2%) | 128<br>(34.4%) |
| 6 | I think vaccination will help us stop the spread of COVID-19. | 3.80<br>(1.10) | 19<br>(5.1%) | 29<br>(7.8%) | 68<br>(18.3%) | 149<br>(40.1%) | 107<br>(28.8%) |
| 7 | I think I'm not informed enough about vaccines. | 2.70<br>(1.27) | 75<br>(20.2%) | 107<br>(28.8%) | 81<br>(21.8%) | 71<br>(19.1%) | 38<br>(10.2%) |
| 8 | Are you hesitant to take the COVID-19 vaccine? | 2.89<br>(1.35) | 71<br>(19.1%) | 87<br>(23.4%) | 89<br>(23.9%) | 63<br>(16.9%) | 62<br>(16.7%) |
| 9 | Should the vaccine be mandatory for healthcare workers? | 3.08<br>(1.52) | 86<br>(23.1%) | 64<br>(17.2%) | 50<br>(13.4%) | 78<br>(21.0%) | 94<br>(25.3%) |
| 10 | Should the vaccine be mandatory for all South Africans? | 2.83<br>(1.45) | 98<br>(26.3%) | 67<br>(18.0%) | 71<br>(19.1%) | 72<br>(19.4%) | 64<br>(17.2%) |

m – mean; SD – standard deviation

### Association between sociodemographic characteristics and HCWs KAP

The study examined the knowledge, attitudes, and perceptions of HCWs towards voluntary and mandatory COVID-19 vaccinations, focusing on sex and age differences (Table 5). Analysis revealed that sex did not significantly impact attitudes and perceptions, knowledge, or vaccine hesitancy, indicating similar levels of understanding and acceptance between male and female HCWs. The ANOVA results showed no significant sex-based differences in attitudes (F = 2.335, p = 0.127), knowledge (F = 0.002, p = 0.962), or hesitancy (F = 0.988, p = 0.321).

**Table 5.** ANOVA Results for Mean Groups Based on Gender and Age.

| Variable | Source | Sum of Squares | Mean Square | F | Sig. |
| --- | --- | --- | --- | --- | --- |
| **Attitudes and Perceptions** | Gender | 1.084 | 1.084 | 2.335 | 0.127 |
|  | Age (Between Groups) | 7.606 | 2.535 | 5.629 | 0.001 |
|  | Age (Within Groups) | 165.304 | 0.450 |  |  |
| **Knowledge** | Gender | 0.001 | 0.001 | 0.002 | 0.962 |
|  | Age (Between Groups) | 1.956 | 0.652 | 2.005 | 0.113 |
|  | Age (Within Groups) | 119.338 | 0.325 |  |  |
| **Vaccine Hesitancy** | Gender | 0.660 | 0.660 | 0.988 | 0.321 |
|  | Age (Between Groups) | 3.856 | 1.285 | 1.941 | 0.123 |
|  | Age (Within Groups) | 243.086 | 0.662 |  |  |

Conversely, age significantly influenced attitudes and perceptions (F = 5.629, p = 0.001), suggesting that older HCWs might have different views than their younger counterparts. While the knowledge about COVID-19 vaccinations showed a trend towards significance with age (F = 2.005, p = 0.113), it was not statistically significant, implying potential age-related differences in understanding. Vaccine hesitancy was not significantly affected by age (F = 1.941, p = 0.123), indicating that reluctance to vaccinate was consistent across different age groups (Table 5). These findings suggest that educational programmes could be tailored to address specific concerns across age groups to enhance vaccine acceptance.

### Factors Influencing COVID-19 Vaccination Hesitancy: Summary of Exploratory Factor Analysis

The study explored factors influencing COVID-19 vaccination hesitancy among respondents using exploratory factor analysis (EFA) on Likert scale statements (Table 6). The Kaiser-Meyer-Olkin (KMO) measure confirmed sampling adequacy with a value of 0.755, and Bartlett’s test of sphericity was significant, indicating suitable conditions for factor analysis (χ^2^ = 483.602, df = 15, p < 0.001). A single factor was identified, explaining 34.4% of the variance, with prominent loadings on fear of side effects (0.679) and insufficient knowledge (0.671), highlighting key contributors to vaccine hesitancy. Communalities analysis showed that concerns about eligibility, knowledge, and side effects saw increased explanation post-extraction, while cost and religious beliefs showed lesser impacts.

**Table 6.** Factor Analysis Results for factors influencing COVID-19 vaccine hesitancy.

| Analysis Results | Values | Variables | Factor Loadings | Communalities |
| --- | --- | --- | --- | --- |
| KMO Measure | 0.755 | Fear of Side Effects | 0.679 | Initial: 0.383 |
| Bartlett's Test ($\chi^2$) | 483.602 (p < 0.001) | Insufficient Knowledge | 0.671 | Extraction: 0.461 |
| Eigenvalue for Factor 1 | 2.064 | Other Reasons | 0.626 | Initial: 0.338 |
| % of Variance Explained | 34.4% | Ineligibility Perception | 0.561 | Extraction: 0.391 |
|  |  | Religious Beliefs | 0.509 | Initial: 0.266 |
|  |  | Cost Concerns | 0.432 | Extraction: 0.260 |

## Discussion

The inclusion of individuals aged 26 to 40 years in this study aligns with Karlsson and Antfolk’s (2021) research, which, although not specifically focusing on age distribution, emphasizes the importance of understanding vaccination confidence and behaviour across different age groups within the healthcare profession.^11^ The gender distribution among the 372 HCWs at the Nursing College, where 81.2% were female and 18.8% male, reveals a significant gender imbalance. This finding is consistent with global trends identified by Dror et al. (2020), who noted that women often exhibit greater vaccine reluctance. Understanding these gender-specific attitudes is crucial for contextualising our analysis of vaccine uptake.^12^

The marital status distribution among these HCWs shows a predominance of single respondents (67.9%). This observation supports Karlsson and Antfolk’s (2021) assertion on the importance of considering demographic variables like marital status when examining vaccination confidence and behaviour.^11^ Additionally, the study population was comprised predominantly of students (92.5%), with a small percentage in administrative (1.9%) and academic roles (5.6%). This distribution reflects global patterns highlighted by Dror et al. (2020),^12^ which suggest that educational attainment and access to information influence vaccine willingness. These findings underscore the importance of understanding vaccine attitudes among younger demographics, such as students.

The current study revealed variability in COVID-19 vaccination uptake. While most respondents were fully vaccinated without boosters, others received both full vaccination and boosters. This high vaccination rate at the Nursing College indicates effective disease prevention. The data shows that most HCWs were vaccinated voluntarily, with only a few under mandate, demonstrating that their rights were respected while they actively chose to protect their colleagues and patients. Nonetheless, the debate on compulsory vaccination remains complex and warrants further consideration. The effectiveness of the COVID-19 vaccine is evident, as most respondents had not been previously infected, and only a few contracted the virus despite being vaccinated. Preferences for receiving health-related information via television and radio reflect the need for accessible communication, especially for older adults who may not use modern digital media extensively.

Islam et al. (2021) conducted a community survey evaluating attitudes towards COVID-19 vaccinations, providing essential context for understanding healthcare professionals’ acceptance.^13^ Their findings emphasise the importance of considering diverse perspectives and experiences, which helps clarify the complex landscape of vaccination uptake and hesitancy. Our study reveals that most HCWs at the Nursing College were vaccinated, predominantly voluntarily, supporting the acceptance of COVID-19 vaccinations. This result rejects the null hypothesis (H0) and confirms the alternate hypothesis (H1) that HCWs generally accept COVID-19 vaccinations, whether mandatory or voluntary.

The research indicates that COVID-19 vaccinations are considered highly effective. Although a few individuals were infected post-vaccination, this outcome supports the vaccine’s safety and efficacy. The observed discrepancies in post-vaccination preventative measures highlight the need for clear communication to address ongoing preventive behaviours. Effective strategies must emphasise the continued importance of preventive measures even after vaccination. Poor communication about the efficacy of COVID-19 vaccinations in preventing severe disease reflects the need for better information dissemination. The lack of consensus on vaccination status limitations underscores the complexity of balancing individual liberties with public health measures.

Awareness of vaccine-related resources varied among respondents, highlighting the necessity of clear, accessible communication. Concerns about potential adverse effects emphasise the need for effective communication strategies to address these issues and reinforce the safety of vaccines. Divergent opinions on vaccinating individuals with pre-existing conditions and pregnant women stress the importance of targeted outreach strategies. The study reveals that adverse effects of COVID-19 vaccination are generally mild and short-lived. However, concerns about potential long-term effects necessitate skilful communication to build public confidence in vaccine safety. The correlation between knowledge, attitudes, and vaccine acceptance indicates that understanding these factors is crucial for enhancing vaccine uptake.

The study also reflects varying degrees of apprehension about the COVID-19 pandemic, with a majority expressing significant concern. Addressing the psychological impacts of the pandemic and incorporating mental health considerations into public health messaging is crucial. The survey indicates broad support for the effectiveness and safety of the vaccine. These findings underscore the urgency of vaccination and the need for focused communication strategies that highlight its importance. The study emphasises the role of community-based communication and social media in promoting vaccination. Encouraging family and neighbours to get vaccinated highlights the impact of social influencers on vaccine perceptions.

### The public health implications of this study

The significant adoption of COVID-19 immunisations among HCWs, mostly via voluntary methods, highlights the need to encourage volunteer vaccination initiatives. It demonstrates the readiness of HCWs to safeguard themselves, their peers and patients against the illness. This is important for other vaccines, such as the influenza vaccine. Public health initiatives should continue to highlight the advantages of vaccination and address any lingering doubts or misunderstandings to sustain high acceptance rates. The research emphasises the need to use efficient communication tactics to influence vaccination attitudes and behaviours.

Although there has been a generally high acceptance of COVID-19 immunisations, the research highlights that some HCWs still have reservations and are hesitant to be vaccinated. These worries may arise from various sources, such as disinformation, apprehension about negative impacts and ambiguity over long-term repercussions. Public health interventions should prioritise resolving vaccination hesitancy among HCWs via focused educational programmes, honest communication and personalised methods that recognise and tackle unique concerns. The research emphasises the need to maintain COVID-19 preventive measures, even among persons who have been vaccinated.

The discourse pertaining to compulsory vaccination elicits significant ethical debates. Although most HCWs in the survey chose to get vaccinated willingly, there were differing views on whether vaccination should be mandated. Public health authorities must thoroughly evaluate ethical principles, individual rights and the possible ramifications of compulsory vaccination programmes on healthcare workers’ autonomy and faith in healthcare systems. When considering whether to require vaccination, it is important to follow ethical guidelines and engage in effective communication to address any concerns and create better understanding. The research emphasises the need for customised strategies for vaccine outreach, specifically targeting vulnerable groups such as pregnant women and persons with pre-existing health issues. To guarantee fair distribution and acceptance of vaccines across all demographic groups, it is important for public health campaigns to prioritise focused communication, easy access to information, and healthcare settings that provide support.

### Limitations of the Study

Although the research achieved a high participation rate, sampling bias is possible, especially if some HCWs were more inclined to participate than others. This has the potential to restrict the applicability of the results to a wider community of HCWs. The research depends on data that individuals describe about themselves, which might be influenced by memory bias or a tendency to provide socially desirable responses. Respondents may tend to exaggerate their favourable views towards vaccination or downplay their reluctance, which might result in possible mistakes in the findings. The study’s cross-sectional design hinders demonstrating causal correlations between variables. Although the research offers useful insights regarding vaccination attitudes and behaviours at a single moment, longitudinal studies would be necessary to evaluate changes over time and the effects of treatments.

The research was conducted at a solitary nursing institution, restricting the applicability of the results to other healthcare settings or areas. Healthcare infrastructure, cultural norms and vaccine delivery tactics in various situations may have varying effects on vaccination dynamics.

## Conclusion

This study demonstrates that while there is general support for COVID-19 vaccination, concerns about vaccine safety and long-term effects persist. Addressing these concerns through targeted communication and education is essential for increasing vaccine acceptance and utilisation. The diverse attitudes and perceptions revealed by the study underscore the need for a comprehensive strategy to address vaccine hesitancy, with a focus on safety and knowledge deficits.

## Data Availability

Data is available upon request and within the prescripts of the Protection 525 of Personal Information Act (POPIAct).

## Supporting information

**S1 Appendix. Study questionnaire**

## Acknowledgments

Special thanks to the Nursing college management for permitting the study and to the participants for their involvement in realising the goals of the research.

